# Genetic Variations in TrkB.T1 Isoform and Their Association with Somatic and Psychological Symptoms in Individuals with IBS

**DOI:** 10.1101/2023.09.14.23295434

**Authors:** H. Hong, E. Mocci, K. Kamp, S. Zhu, K.C. Cain, R. L. Burr, J. Perry, M. M Heitkemper, K. R. Weaver-Toedtman, S.G Dorsey

**Author notes:** Corresponding Author: Susan G. Dorsey, PhD RN FAAN University of Maryland School of Nursing 655 W Lombard St, Baltimore, MD 21201, USA. These authors contributed equally to this manuscript (H. Hong, E. Mocci).

## Abstract

Irritable bowel syndrome (IBS), a disorder of gut-brain interaction, is often comorbid with somatic pain and psychological disorders. Dysregulated signaling of brain-derived neurotrophic factor (BDNF) and its receptor, tropomyosin-related kinase B (TrkB), has been implicated in somatic-psychological symptoms in individuals with IBS. Thus, we investigated the association of 10 single nucleotide polymorphisms (SNPs) in the regulatory 3’ untranslated region (UTR) of *NTRK2* (TrkB) kinase domain-deficient truncated isoform (TrkB.T1) and the *BDNF* Val66Met SNP with somatic and psychological symptoms and quality of life in a U.S. cohort (IBS n=464; healthy controls n=156). We found that the homozygous recessive genotype (G/G) of rs2013566 in individuals with IBS is associated with worsened somatic symptoms, including headache, back pain, joint pain, muscle pain, and somatization as well as diminished sleep quality, energy level and overall quality of life. Validation using U.K. BioBank (UKBB) data confirmed the association of rs2013566 with increased likelihood of headache. Several SNPs (rs1627784, rs1624327, rs1147198) showed significant associations with muscle pain in our U.S. cohort. Notably, these SNPs are predominantly located in H3K4Me1-enriched regions, suggesting their enhancer and/or transcription regulation potential. Together, our findings suggest that genetic variation within the 3’UTR region of the TrkB.T1 isoform may contribute to comorbid conditions in individuals with IBS, resulting in a spectrum of somatic and psychological symptoms that may influence their quality of life. These findings advance our understanding of the genetic interaction between BDNF/TrkB pathways and somatic-psychological symptoms in IBS, highlighting the importance of further exploring this interaction for potential clinical applications.

## Introduction

Irritable bowel syndrome (IBS) is a disorder of gut-brain interaction characterized by altered bowel habits and chronic abdominal pain, resulting from the complex interaction between the gut and the brain [1, 2]. Individuals with IBS often experience extraintestinal symptoms such as backache, headache, joint pain, fatigue, and sleep disturbances [3]. Furthermore, they have a higher prevalence of neurological pain conditions such as fibromyalgia and migraine compared to individuals without IBS [4]. Psychological symptoms and disorders are also commonly associated with IBS, with individuals having a three-fold higher risk of anxiety or depression compared to healthy individuals [5]. Numerous mechanisms have been explored to understand the symptomatology of IBS, including abnormalities in serotonin and bile acid metabolism, altered intestinal permeability, immune activation, disturbances in the intestinal microbiota, and changes in brain function [6]. Genetic factors have been shown to play a significant role in IBS, as evidenced by studies utilizing epigenetic, genome-wide association, and candidate gene association analyses [7–9]. However, the identification of specific genes and variants associated with both gastrointestinal and extraintestinal symptoms in individuals with IBS remains elusive.

Brain-derived neurotrophic factor (BDNF) signaling has emerged as a pivotal player in determining diverse somatic pain [10–12] and psychological disorders [13, 14]. BDNF belongs to the neurotrophin family of growth factors and plays a crucial role in neuronal development, survival, and plasticity within the central and peripheral nervous systems [15–17]. BDNF transmits intracellular signaling through binding to the extracellular domain of the tropomyosin-related kinase B (TrkB) receptor, which is encoded by the neurotrophic receptor tyrosine kinase 2 (*NTRK2*) gene [18]. The *NTRK2* locus encodes multiple receptor isoforms involved in different functional activities in BDNF signaling. The primary isoform of the TrkB receptor, known as full-length TrkB (TrkB.FL), possesses a C-terminal tyrosine kinase domain (exon 20-24) that mediates several canonical signaling pathways, including phosphoinositide 3-kinase-protein kinase B (PI3K/Akt), mitogen-activated protein kinases (MAPK), and phospholipase C-γ (PLCγ) signaling [19]. In contrast, the C-terminal truncated isoform, TrkB.T1, lacks the intracellular kinase domain, resulting in limited canonical or alternative BDNF signaling [20]. In both human and animal models, TrkB.T1-mediated non-canonical signaling and differential expression of TrkB.T1 have been shown to be associated with the increased risk of neuropathic pain [21–23] and mood and psychological disorders [24–26]. Therefore, the function of TrkB.T1 in modifying BDNF signaling highlights its significance in the pathobiology of various health conditions, emphasizing its potential role in critical somatic and psychological manifestations.

Understanding the genetic effects of the TrkB.T1 isoform on somatic and psychological symptoms in IBS could be valuable in elucidating underlying mechanisms and improving diagnostic and therapeutic approaches. The TrkB.T1 isoform is generated via alternative splicing, specifically through utilization of exon 16. This exon contains regulatory elements such as the H3K4me1 histone mark and polyadenylation sites in its long (>500 nucleotide) 3’ untranslated region (UTR), a region that is essential for gene expression and functional regulation [24, 27–29]. Thus, we hypothesized that genetic variations in the 3’UTR of the TrkB.T1 isoform may regulate BDNF/TrkB signaling mechanisms, influencing the manifestation of somatic and psychological symptoms commonly observed in individuals with IBS. Therefore, this study aims to investigate the associations of single nucleotide polymorphisms (SNPs) in *BDNF* and the TrkB-T1 isoform specific locus of *NTRK2* with somatic and psychological symptoms in IBS. We specifically focused on genotypes for 14 SNPs, including 13 located in the regulatory 3’UTR region of the TrkB-T1 isoform, and the well-established functional SNP *BDNF* Val66Met (rs6265), known to affect the release of BDNF protein [30]. By analyzing the genotypic associations of these SNPs with symptoms in three domains— (i) abdominal pain and other somatic symptoms, (ii) psychological symptoms, and (iii) quality of life—we aimed to gain a comprehensive understanding of their genetic effects on IBS-related symptomatology. Furthermore, the associations between symptoms and genotypes were externally validated using the United Kingdom BioBank (UKBB) data, enhancing the reliability of our findings.

## Materials and Methods

### Study Participants

This study examined participant data across five independent research studies conducted at a research center in the Pacific Northwest of the United States (U.S.) (Figure 1). Among these studies, three were observational and focused on understanding the mechanisms of IBS. They recruited women with IBS and women without the condition, who served as healthy controls (HCs) within the age range of 18-50 [31–33]. The remaining two studies were randomized controlled trials that investigated a comprehensive self-management intervention. They recruited both men and women with IBS within the age range of 18-70 [34, 35]. For the current analysis, only the baseline data prior to randomization from the intervention studies was utilized. Detailed information about the complete study protocols can be found elsewhere [31–36]. Briefly, participants for all studies were recruited through mailings from a gastroenterology practice and community advertisements. The eligibility criteria were similar across the studies. Individuals with IBS had a confirmed diagnosis for at least 6 months by a healthcare provider and met the Rome III research criteria for IBS. The HCs were required to have no moderate to severe diseases, disorders, or symptoms. Additionally, individuals with a history or current experience of IBS-like symptoms were excluded from the HC group. Baseline blood samples were obtained from all participants to conduct targeted genetic testing.

**Figure 1.**
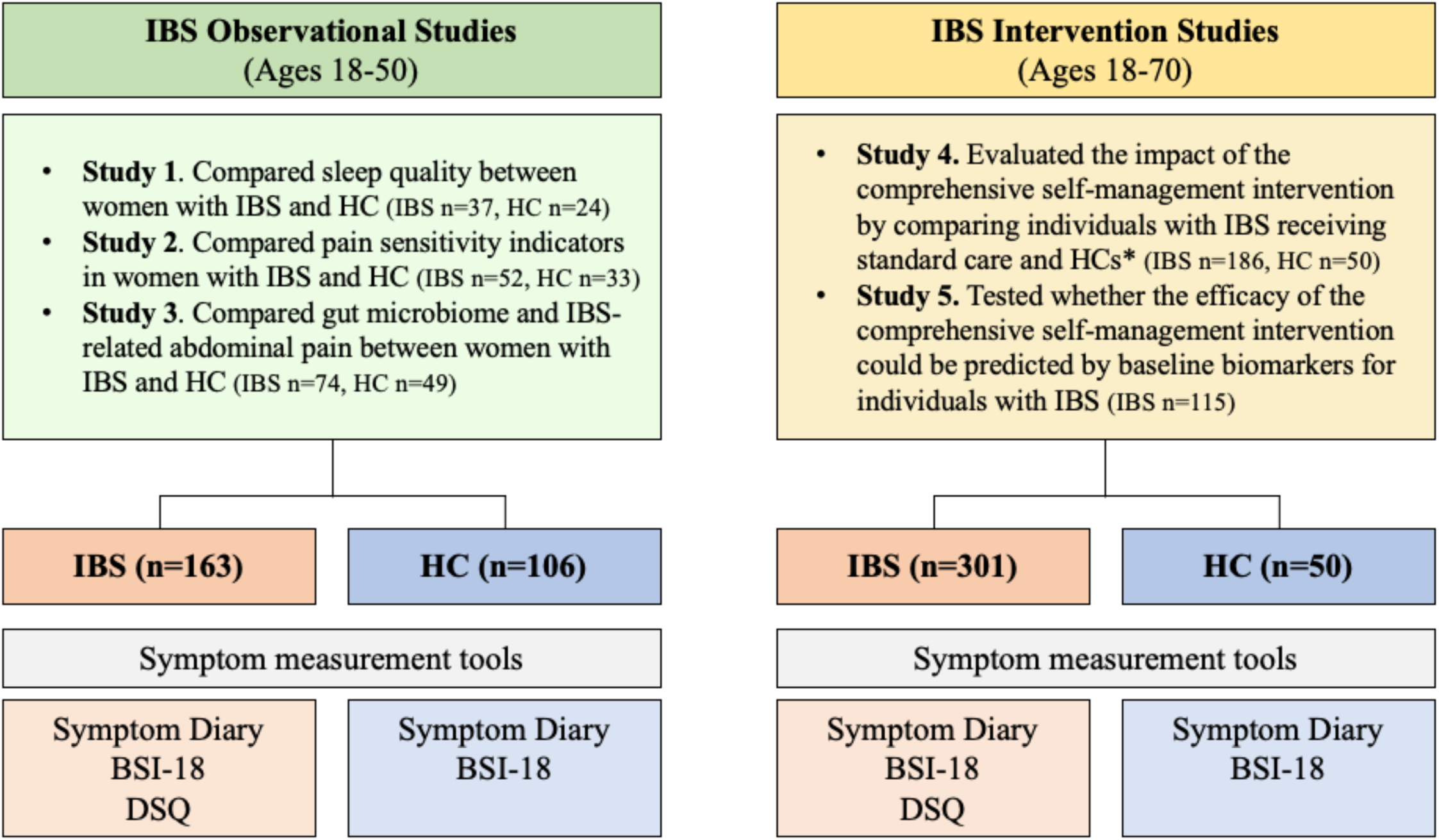
Flow Diagram Illustrating Data Utilization in Five IBS Studies. *HCs were recruited solely for questionnaire and DNA sampling purposes, without any intervention being provided. BSI-18: Brief Symptom Inventory-18, DSQ: Disease Specific Questionnaire, HC: healthy control

### Study Measures

*Demographic survey* collected participant information on self-identified age, race, ethnicity, gender, marital status, employment, education, and family income using a demographic questionnaire.

The *Daily Symptom Diary* was used to record the severity of symptoms in a daily diary. Participants were asked to ‘mark the highest severity that you have experienced with each symptom over the past 24 hours’, with symptoms rated as: not present (0), mild (1), moderate (2), severe (3), and very severe (4). Ratings were averaged for each symptom. Daily symptoms included in this analysis are abdominal pain or discomfort, somatic pain (backache, headache, joint pain, and muscle pain), and psychological symptoms (anxiety, depressed/sad or blue). The *Brief Symptom Inventory (BSI-18)* [37] was used to measure psychological distress, including symptoms of anxiety, depression, and somatization. Participants responded on a scale from (0) not at all to (4) extremely. Internal consistency of symptom dimensions is reported as .71 to .85, and test-retest reliability from .68 to .91 [38, 39].

The IBS-related quality of life was evaluated with the *Disease Specific Questionnaire (DSQ)* [40]. Participants reported the impact of IBS symptoms on their well-being over the past four weeks. Scores range from 0 to 100 for each subscale: emotional well-being, mental health, sleep, energy, physical functioning, food, social interactions, physical role, and sexual relations. Higher scores on these subscales indicate a higher quality of life. The DSQ was administered solely to the IBS group since it is a specific tool for measuring quality of life in individuals with this condition.

### Biospecimen Collection and DNA Extraction

Biospecimens were collected per protocol of the above referenced investigations. Genomic DNA (gDNA) was extracted from either frozen buffy coat preparations [41] or fresh whole blood using Qiagen DNeasy Blood & Tissue kits (Qiagen, Valencia, CA) or Puregene DNA Purification kits (Gentra Systems Inc., Minneapolis, MD). Samples were stored at -80°C until shipped overnight on dry ice to the Institute for Genome Sciences at the University of Maryland, Baltimore.

### SNP Genotyping and Quality Assurance

Genotyping was performed using TaqMan® SNP Genotyping Assays (Applied Biosystems, Life Technologies Corporation), which include two sequence-specific primers for amplification of sequences containing the SNPs of interest, and two allele-specific TaqMan™ probes for Allele 1 and Allele 2. The samples were thawed, and gDNA was quantitated. A reaction mix containing TaqMan® Genotyping Assay (20X), TaqMan® Genotyping Master Mix, and nuclease-free water was prepared. The reaction mix and samples were aliquoted into 96-well plates, organized by study. The plates were then run on QuantStudioX Real-Time PCR System, with analysis of genotyping experiments performed using QuantStudio™ Design and Analysis desktop Software. Genotyping was performed using the GRCh38 reference genome as the basis for variant calling and allele identification. Detailed information on the 14 genotyped SNPs, including the closest gene, function, frequency, and minor allele frequency (MAF) in our study and the European population of the 1000 Phase 3 Genome Project [42], is provided in Supplemental Table 1. To assess the quality of the genotyping, we controlled the genotypes cluster for each SNP. Two SNPs, rs2013566 and rs3654, located in the 3’UTR of *NTRK2* gene, failed genotyping in one out of the five studies, and therefore contained 20% less samples for these SNPs. The remaining SNPs were successfully genotyped in all studies with an average number of missing genotyped samples equal to 5.5 (range 3-11). As an additional quality control measure, considering our sample’s 75% Caucasian ancestry, we compared the MAF of all 14 SNPs with their MAF in the European population of the 1000 Phase 3 Genome Project. We observed a high correlation between the computed allele frequencies of the SNPs in our sample and those of the 1000 Genomes dataset. Two SNPs, rs121434633 and rs74356179, were excluded from the analysis due to their extremely low MAF values in our samples (0.000 and 0.002, respectively; Supplemental Table 1). Next, we measured pairwise squared correlation (r^2^) linkage disequilibrium (LD) among the SNPs in our samples using Haploview [43]. A strong correlation (r^2^ = 0.9) was observed between rs45596934 and rs138535351, leading us to exclude rs45596934 due to its higher number of missing genotypes compared to rs138535351. None of the remaining SNPs in the *NTRK2* gene exhibited a significant correlation (r^2^ < 0.6, Figure 2). In conclusion, after conducting a thorough quality check of the 14 genotyped SNPs, we retained 11 SNPs for downstream analyses, with 10 of them located in the *NTRK2* gene and 1 in the *BDNF* gene. The SNP locations within the *NTRK2* gene and their association with the most common transcripts are shown in Figure 2.

**Figure 2.**
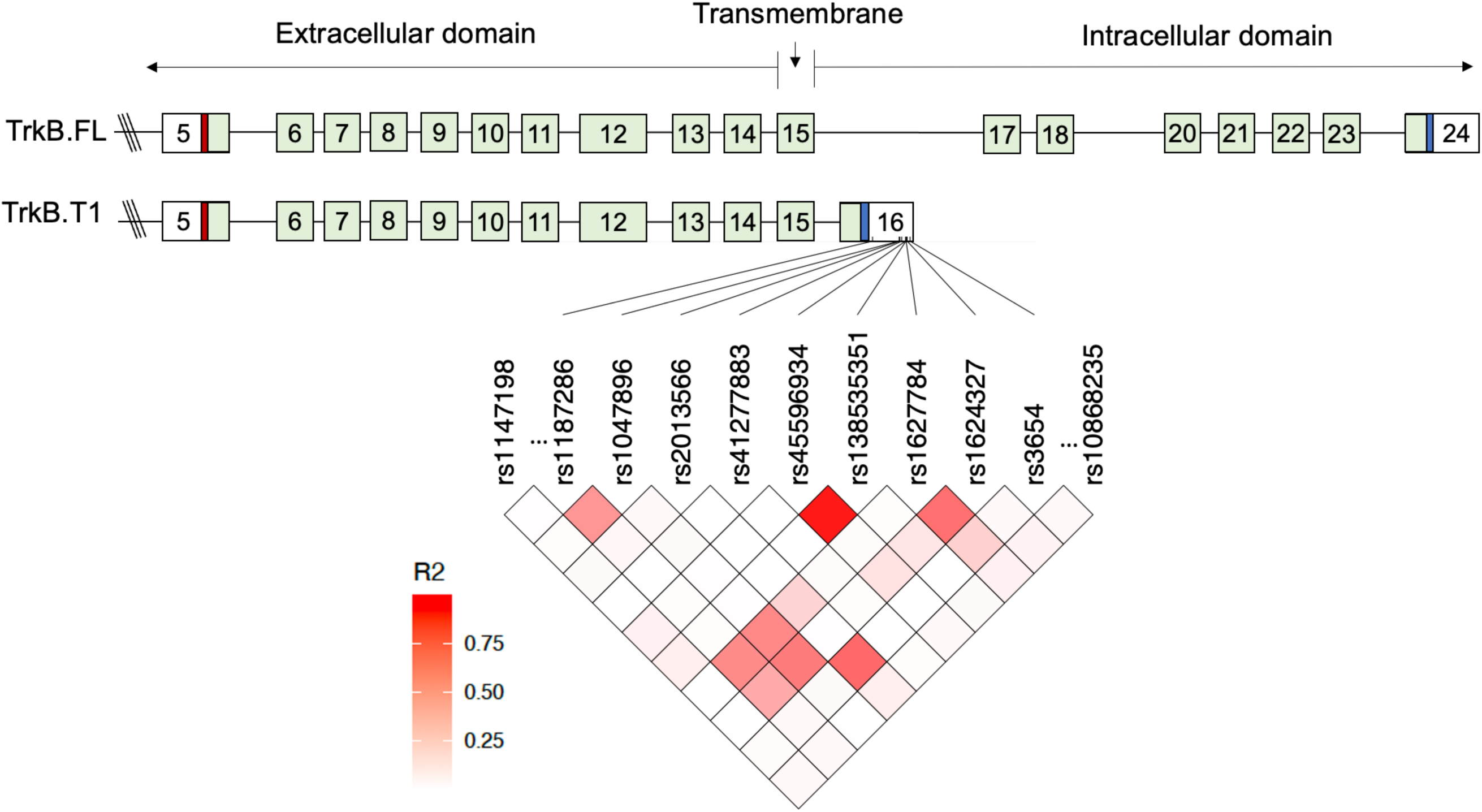
TrkB.T1 Isoform and SNPs in 3’ untranslated region (UTR). ***(Top) Structure of the TrkB isoforms***. Protein coding regions are shown as light green boxes and exons are numbered. 5′ and 3′ UTR regions are shown as empty boxes. The location of translational start codon (red) and stop codon (blue) are shown as thin boxes. TrkB.FL contains an intracellular C-terminal tyrosine kinase domain (exon 20-24) and is involved in various signaling pathways, including PI3K/Akt, MAPK, and PLCγ signaling. The truncated isoform, TrkB.T1, lacks the intracellular kinase domain due to the presence of exon 16, which contains 33 nucleotides that encode a unique intracellular 11 amino acid domain, a stop codon, and a unique 3’ UTR sequence with multiple polyadenylation sites. ***(Bottom) 3’ UTR SNPs in TrkB.T1 Isoform and LD Heatmap.*** The bottom figure illustrates the pass-filter 11 SNPs located within the 3’ UTR of the TrkB.T1 isoform, where our candidate SNPs are positioned. The heatmap represents R^2^ linkage disequilibrium (LD) for the SNPs in this region. Based on the strong correlation (R^2^= 0.9) observed between rs45596934 and rs138535351, rs45596934 was omitted from the analysis due to a relatively higher number of missing genotypes than rs138535351. Note that rs1147198 and rs10868235 are situated in an upstream intergenic region and a downstream intronic region, respectively, outside of exon 16. Additionally, rs6265 is located in the *BDNF* gene on a different chromosome (not shown). The isoform structure in the top figure has been modified from Figure 1 in Timmusk et al. (2010) with permission from John Wiley and Sons.

### Data analyses

The participant data from the five studies were merged, and demographic characteristics were compared between participants with IBS and HCs using t-tests and chi-square tests, with a significance level set at p < 0.05, using R studio [44]. Logistic regression analysis was initially conducted with 11 SNPs to test for their association with IBS status (IBS n = 464, HC n = 156), controlling for age, sex, and study design. Our primary analysis of the relationship between *NTRK2* and *BDNF* SNPs and symptoms was conducted using an additive linear regression model, considering that symptoms were measured as continuous variables. To investigate potential dominance or recessive effects, we further employed a genotypic modifier, which in addition to testing the additive (ADD) model (number of minor alleles coded as 0, 1, or 2) assessed significant deviation from dominance (DOMDEV) by comparing heterozygote genotypes (coded as 1) to homozygotes (coded as 0). Finally, the genotypic modifier implemented a joint test (GENO_2DF) to evaluate the combined significance of additive and dominance-deviation terms [45]. The significant effects observed solely in the ADD model indicate a linear dose-dependent relationship between the number of minor alleles and the trait. Additionally, the consistent and significant findings in the DOMDEV and GENO_2DF models, aligning with the ADD model, suggest a potential dominance effect. Moreover, when all three tests are significant and ADD and DOMDEV have effects with opposite directions, it suggests a recessive effect. To maximize the robustness of our findings, we do not consider associations as significant if they are significant only in the DOMDEV and GENO_2DF models but not in the ADD model. To ensure accurate estimation of effect sizes, significant genetic associations are determined based on the criterion that all three genotypes (homozygous dominant, heterozygous, and homozygous recessive) should be represented by a minimum of two individuals. Both logistic and linear regression analyses were performed using PLINK (Version 2) [46, 47]. To address potential confounding factors arising from independent study recruitment strategies, all regression models were adjusted for age, sex, and study design. To account for the increased risk of false positive results due to multiple testing, we employed the Benjamini-Hochberg procedure to estimate and control the false discovery rate (FDR).

### Validation analyses

To ensure the reliability of our results, we conducted external validation of the associations between 11 SNPs and abdominal/somatic and psychological symptoms. This validation was performed using publicly available databases, namely the ES200kUKB and ImpUKB datasets, which were curated using the Omics Analysis, Search & Information System (OASIS) [48]. The ES200kUKB dataset consisted of participants from the UKBB who underwent whole exome sequencing, enabling comprehensive identification of genetic variants [49]. The ImpUKB dataset represented imputed genotypes of UKBB participants based on the Haplotype Reference Consortium (HRC) and UK10K haplotype resource, allowing accurate imputation of missing genotypes [50]. Using International Classification of Diseases, 10^th^ revision (ICD-10) codes for IBS, pain, and psychiatric diagnoses (Supplemental Table 2), we identified individuals with IBS and symptoms aligning closely with those in our primary investigation. This enabled us to differentiate cases (IBS individuals with one or more comorbid somatic and psychological symptoms based on ICD-10 codes) from controls (IBS individuals without comorbid symptoms based on ICD-10 codes). Of 200,643 individuals in ES200kUKB dataset, 3,060 were diagnosed with IBS according to ICD-10 codes. Similarly, the ImpUKB dataset included 487,409, and 7,568 individuals diagnosed with IBS. Logistic regression using an additive genetic model was used to test the associations between the SNPs and 9 ICD-10 diagnoses, specifically comparing individuals who had comorbid diagnoses of IBS with other somatic symptoms (backache, headache, joint pain, muscle pain, impaired sleep, decreased energy, and/or somatization) and psychological diagnoses (anxiety, depression) to HCs. We applied MAF greater than or equal to 0.005 as criteria to select SNPs and observed 72 associations between the 11 SNPs and 9 symptom traits in ES200kUKB, and 99 associations in ImpUKB.

## Results

### Demographics and IBS-related symptom characteristics

Genotypes of 11 SNPs were analyzed in 620 participants in the U.S., comprising 464 individuals with IBS and 156 HCs recruited from five independent studies (Figure 1). Table 1 summarizes the baseline characteristics of participants in two groups: IBS Observational Studies (ages 18-50) and IBS Intervention Studies (ages 18-70). In the IBS Observational Studies group, there were 269 participants (IBS n = 163, HC n = 106), with similar mean ages between the groups (p = 0.42). Although the proportion of White individuals was higher among IBS participants compared to HCs, no significant differences were observed in ethnicity, gender, marital status, employment status, education, or family income between the IBS and HC groups. In the IBS Intervention Studies group, there were 351 participants (IBS n = 301, HC n = 50), with similar mean ages (p = 0.62). HCs were recruited for questionnaire and DNA sampling purposes only, no intervention provided. While no significant differences were found in race, ethnicity, gender, education, or employment status, the HC group had a higher proportion of married/partnered individuals (p = 0.002) and lower family income ≤ 50k (p = 0.03) compared to the IBS group. However, the significane of the difference in marital status is more likely to be influenced by the higher amount of missing data in the HC group compared to the IBS group.

**Table 1.** Baseline Sample Characteristics.

| **IBS Observational Studies (ages 18-50)** |  |  |  |  |
| --- | --- | --- | --- | --- |
|  | Total (n=269) | HC (n=106) | IBS (n=163) | *p* value* |
| **Age, Mean (SD)** | 28.28 (7.03) | 27.87 (6.32) | 28.54 (7.46) | 0.42 |
| **Race, n (%)** |  |  |  | 0.11 |
| Other | 66 (24.54) | 32 (30.19) | 34 (20.86) |  |
| White | 203 (75.46) | 74 (69.81) | 129 (79.14) |  |
| **Ethnicity, n (%)** |  |  |  | 0.52 |
| Hispanic or Latino | 23 (8.55) | 11 (10.38) | 12 (7.36) |  |
| Not Hispanic or Latino | 246 (91.45) | 95 (89.62) | 151 (92.64) |  |
| **Gender, n (%)** |  |  |  |  |
| Female | 268 (99.63) |  |  |  |
| Missing | 1 (0.37) |  |  |  |
| **Marital status, n (%)** |  |  |  | 0.44 |
| Married or partner | 82 (30.48) | 31 (29.25) | 51 (31.29) |  |
| Not married | 186 (69.14) | 74 (69.81) | 112 (68.71) |  |
| **Employment, n (%)** |  |  |  | 0.98 |
| Full time | 84 (31.23) | 32 (30.19) | 52 (31.90) |  |
| Part time | 81 (30.11) | 33 (31.13) | 48 (29.45) |  |
| Not working | 42 (15.61) | 16 (15.09) | 26 (15.95) |  |
| Missing | 62 (23.05) | 25 (23.58) | 37 (22.70) |  |
| **Education, n (%)** |  |  |  | 0.14 |
| ≤ 12th grade | 21 (7.81) | 5 (4.72) | 16 (9.82) |  |
| Some college | 168 (66.17) | 70 (66.04) | 108 (66.26) |  |
| Master or higher | 67 (24.91) | 31 (29.25) | 36 (22.09) |  |
| Missing | 3 (1.12) |  | 3 (1.84) |  |
| **Family income, n (%)** |  |  |  | 0.45 |
| < 50k | 103 (38.29) | 44 (41.51) | 59 (36.20) |  |
| 50k -100k | 73 (27.14) | 30 (28.30) | 43 (26.38) |  |
| >100k | 42 (15.61) | 12 (11.32) | 30 (26.38) |  |
| Missing | 51 (18.96) | 20 (11.32) | 31 (19.02) |  |
| **IBS Intervention Studies (ages 18-70)** |  |  |  |  |
|  | Total (N=351) | HC (n=50) | IBS (n=301) | *p* value |
| **Age, Mean (SD)** | 42.91 (14.7) | 43.88 (14.8) | 42.74 (14.7) | 0.62 |
| **Race, n (%)** |  |  |  | 1.00 |
| Other | 66 (18.8) | 9 (18) | 57 (18.94) |  |
| White | 285 (81.2) | 41 (82) | 244 (81.06) |  |
| **Ethnicity, n (%)** |  |  |  | 0.13 |
| Hispanic or Latino | 12 (3.42) | 4 (8) | 8 (2.66) |  |
| Not Hispanic or Latino | 339 (96.58) | 46 (92) | 293 (97.34) |  |
| **Gender, n (%)** |  |  |  | 0.87 |
| Female | 294 (83.76) | 41 (82) | 253 (84.05) |  |
| Male | 57 (16.24) | 9 (18) | 48 (15.95) |  |
| **Marital status, n (%)** |  |  |  | 0.002 |
| Married or partner | 149 (42.45) | 20 (40) | 129 (42.86) |  |
| Not married | 200 (56.98) | 28 (56) | 172 (57.14) |  |
| Missing | 2 (0.57) | 2 (4) | 0 |  |
| **Employment, n (%)** |  |  |  | 0.09 |
| Full time | 148 (42.17) | 14 (28) | 134 (44.52) |  |
| Part time | 102 (29.06) | 16 (32) | 86 (28.57) |  |
| Unemployed | 98 (27.92) | 20 (40) | 78 (25.91) |  |
| Missing | 3 (0.85) | 0 | 3 (1.00) |  |
| **Education, n (%)** |  |  |  | 0.98 |
| ≤ 12th grade | 29 (8.26) | 4 (8) | 25 (8.31) |  |
| Some college | 235 (66.95) | 34 (68) | 201 (66.78) |  |
| Master or higher | 86 (24.50) | 12 (24) | 74 (24.58) |  |
| Missing | 1 (0.28) | 0 | 1 (0.33) |  |
| **Family income, n (%)** |  |  |  | 0.03 |
| ≤ 50k | 113 (32.19) | 24 (48) | 89 (21.65) |  |
| 50k -100k | 101 (28.77) | 12 (24) | 89 (21.65) |  |
| >100k | 61 (17.38) | 9 (18) | 52 (17.28) |  |
| Missing | 76 (21.65) | 5 (10) | 71 (23.59) |  |

\**p* values from t-tests or chi-square tests
HC: healthy control, IBS: irritable bowel syndrome, SD: standard deviation, n: number of sample.

Individuals with IBS experience higher levels of somatic pain and psychological distress compared to HCs (Table 2). In the abdominal pain and other somatic symptom domain, IBS participants showed significantly higher mean scores for abdominal pain, back pain, headache, joint pain, muscle pain, and somatization than HCs (p < .0001). Similarly, in the psychological symptom domain, IBS participants exhibited higher mean scores for depression and anxiety than HCs (p < .0001). In the domain of quality of life related to IBS, IBS participants had an overall score of 67.91 (SD = 15.71), indicating a moderate level of quality of life. Together, individuals with IBS experience higher levels of somatic pain and psychological distress compared to HCs.

**Table 2.**
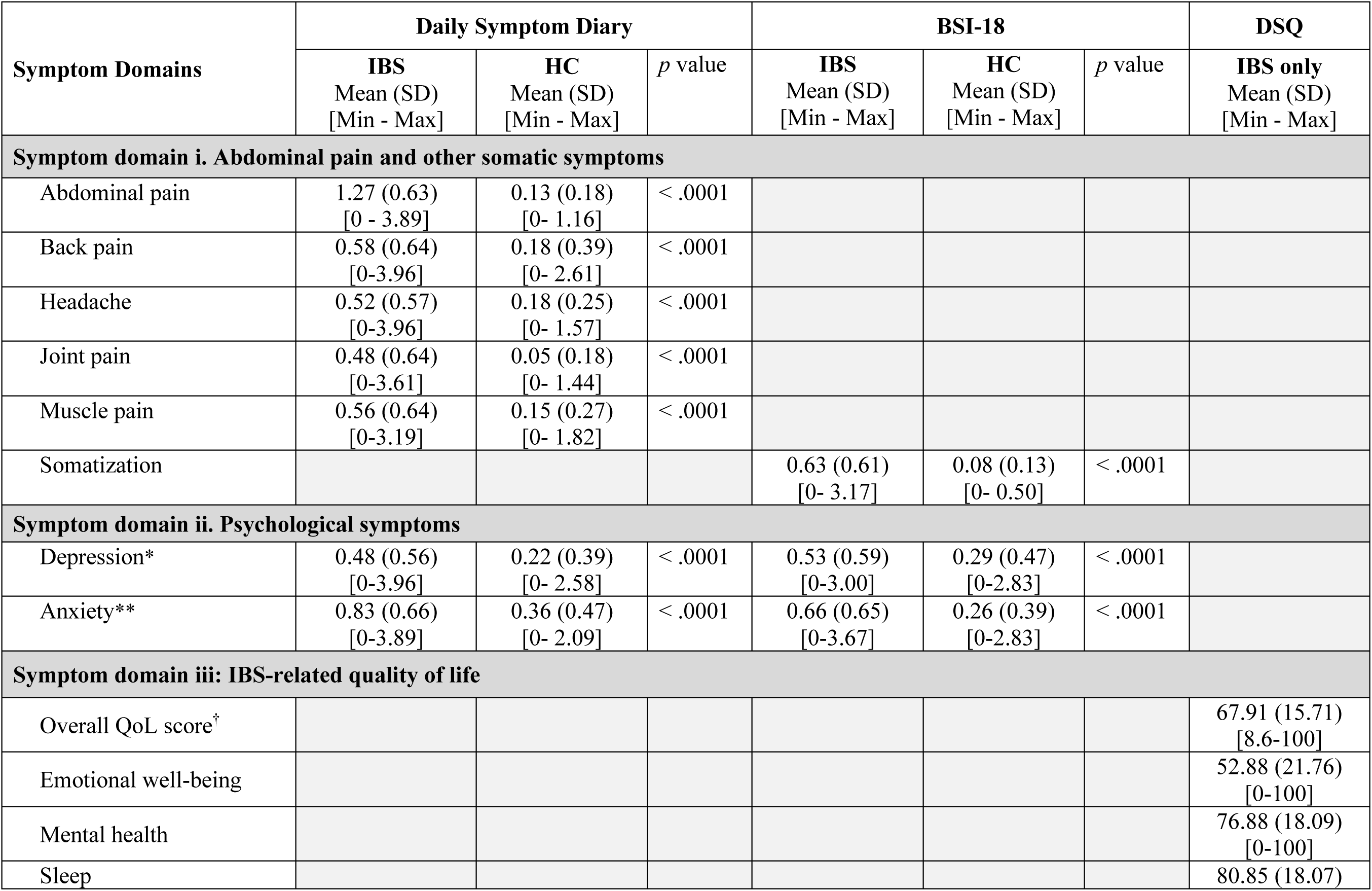
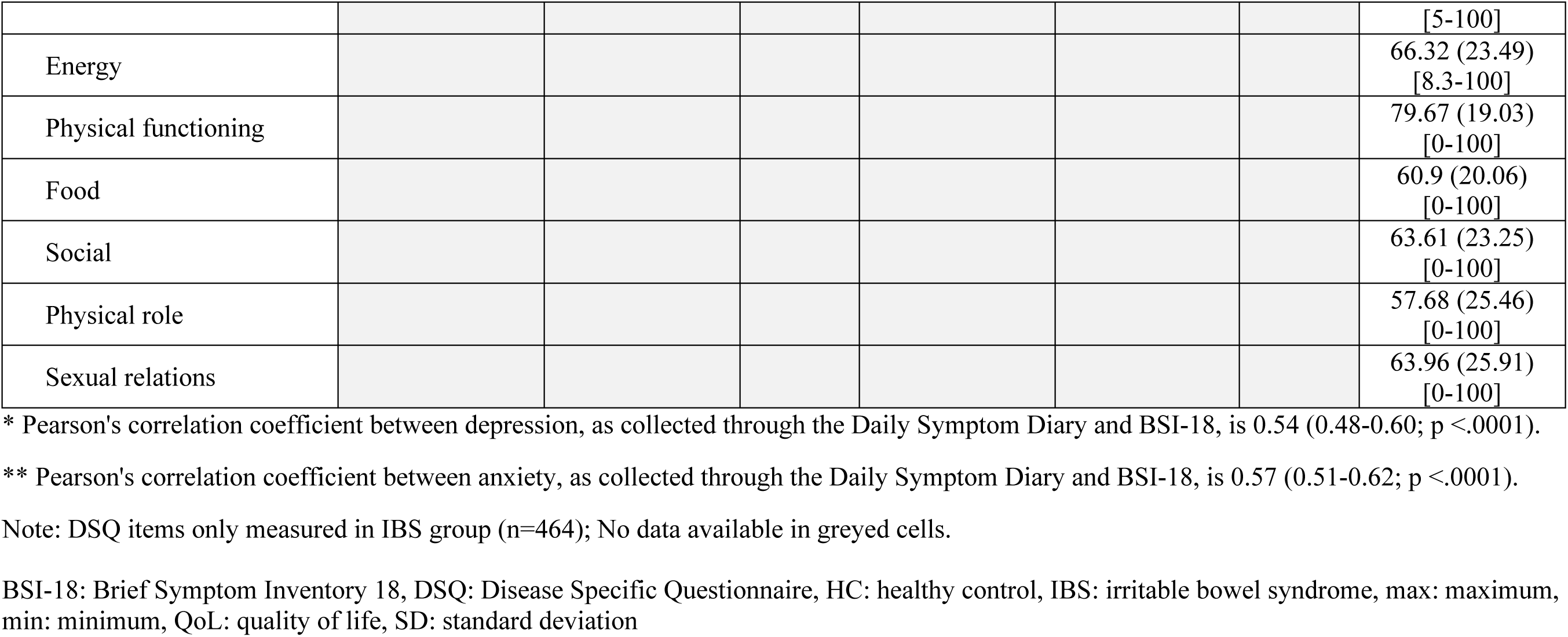
Symptom comparison between IBS and HCs.

### Associations of SNPs in NTRK2 and BDNF genes with IBS phenotype

First, we conducted case-control association analysis using an additive logistic regression model to investigate the potential association between the SNPs and the IBS phenotype. Among the 11 SNPs analyzed, we found that the number of minor alleles at rs41277883 was associated with approximately 3.4 times higher odds of having IBS (95% confidence interval [CI] = 2.30-4.48). However, no significant differences in genotype frequencies were found for the remaining 10 SNPs between individuals with IBS and HCs (Table 3).

**Table 3.** Genetics Association of SNPs and IBS phenotypes.

| SNP ID | Position | REF | ALT | MAF | n | aOR | 95% CI | *p* value |
| --- | --- | --- | --- | --- | --- | --- | --- | --- |
| rs1147198 | 9:84660433 | T | G | 0.266 | 613 | 1.17 | 0.87, 1.47 | 0.31 |
| rs1187286 | 9:84800113 | T | G | 0.244 | 611 | 0.97 | 0.67, 1.26 | 0.82 |
| rs1047896 | 9:84811058 | A | G | 0.187 | 613 | 1.22 | 0.88, 1.57 | 0.25 |
| rs2013566 | 9:84811383 | A | G | 0.113 | 494 | 1.02 | 0.53, 1.51 | 0.93 |
| **rs41277883** | **9:84812145** | **A** | **G** | **0.033** | **611** | **3.39** | **2.30, 4.48** | **0.03** |
| rs138535351 | 9:84813892 | G | A | 0.032 | 615 | 0.83 | 0.12, 1.53 | 0.60 |
| rs1627784 | 9:84813951 | T | C | 0.284 | 612 | 1.08 | 0.78, 1.38 | 0.61 |
| rs1624327 | 9:84814375 | G | A | 0.250 | 607 | 1.05 | 0.75, 1.36 | 0.74 |
| rs3654 | 9:84815576 | T | C | 0.069 | 491 | 0.82 | 0.22, 1.41 | 0.51 |
| rs10868235 | 9:84878840 | C | T | 0.466 | 612 | 0.96 | 0.70, 1.22 | 0.74 |
| rs6265 | 11:27658369 | C | T | 0.200 | 614 | 0.92 | 0.61, 1.23 | 0.60 |

aOR: adjusted odds ratio relative to the alternative allele, ALT: alternate allele, CI: confidence interval, MAF: minor allele frequency, n: number of samples, REF: reference allele

### Uncovering the genetic associations of symptoms in individuals with IBS

Next, we examined the association between 11 SNPs and various quantitative symptom traits, including somatic symptoms, psychological symptoms, and quality of life in individuals with IBS. Using a linear regression model and controlling for age, sex, and study design, we simultaneously tested three genetic models (ADD, DOMDEV, and GENO_2DF; Supplemental Table 3). Notably, the minor allele at rs2013566 demonstrated significant associations with multiple somatic symptom traits, such as back pain, headache, joint pain, muscle pain, and somatization, in the ADD model (p < 0.05; Table 4). Interestingly, these associations were also significant at the DOMDEV and GENO_2DF tests (Supplemental Table 3), indicating the associations between rs2013566 and somatic symptoms follow a recessive pattern (Figure 3a-e). Importantly, contrasting associations were observed within rs2013566 and quality of life domain symptoms. Individuals with a homozygous recessive genotype (G/G) at rs2013566 experienced compromised sleep quality, decreased energy levels, and reduced overall quality of life (p < 0.05, Figure 3f-h). These contrasting associations were supported by DOMDEV and GENO_2DF tests. In the ADD model, the SNP was negatively associated with energy levels and sleep quality, but positively associated in the DOMDEV model (Supplemental Table 3). These findings suggest that individuals with a homozygous recessive genotype at rs2013566 may experience the exacerbation of various somatic symptoms, negatively impacting their quality of life, partly due to poor sleep quality or reduced energy levels. Additionally, three SNPs demonstrated significant associations with specific symptom traits in individuals with IBS, particularly related to muscle pain. For example, the homozygous recessive genotype of rs1147198 showed a positive association with muscle pain (p < 0.05 in three models, Figure 4a), consistent with the findings observed for rs2013566 (Figure 3d). In contrast, the minor alleles of rs1627784 and rs1624327 showed a negative association with muscle pain in the ADD model only (β = -0.12, p = 0.03; β = -0.14, p = 0.03, respectively; Figure 4b, c). Overall, these findings highlight the potential influence of these SNPs on specific symptom traits often associated with IBS, particularly related to somatic pain such as headache and musculoskeletal pain, as well as aspects of quality of life such as sleep quality and energy level.

**Figure 3.**
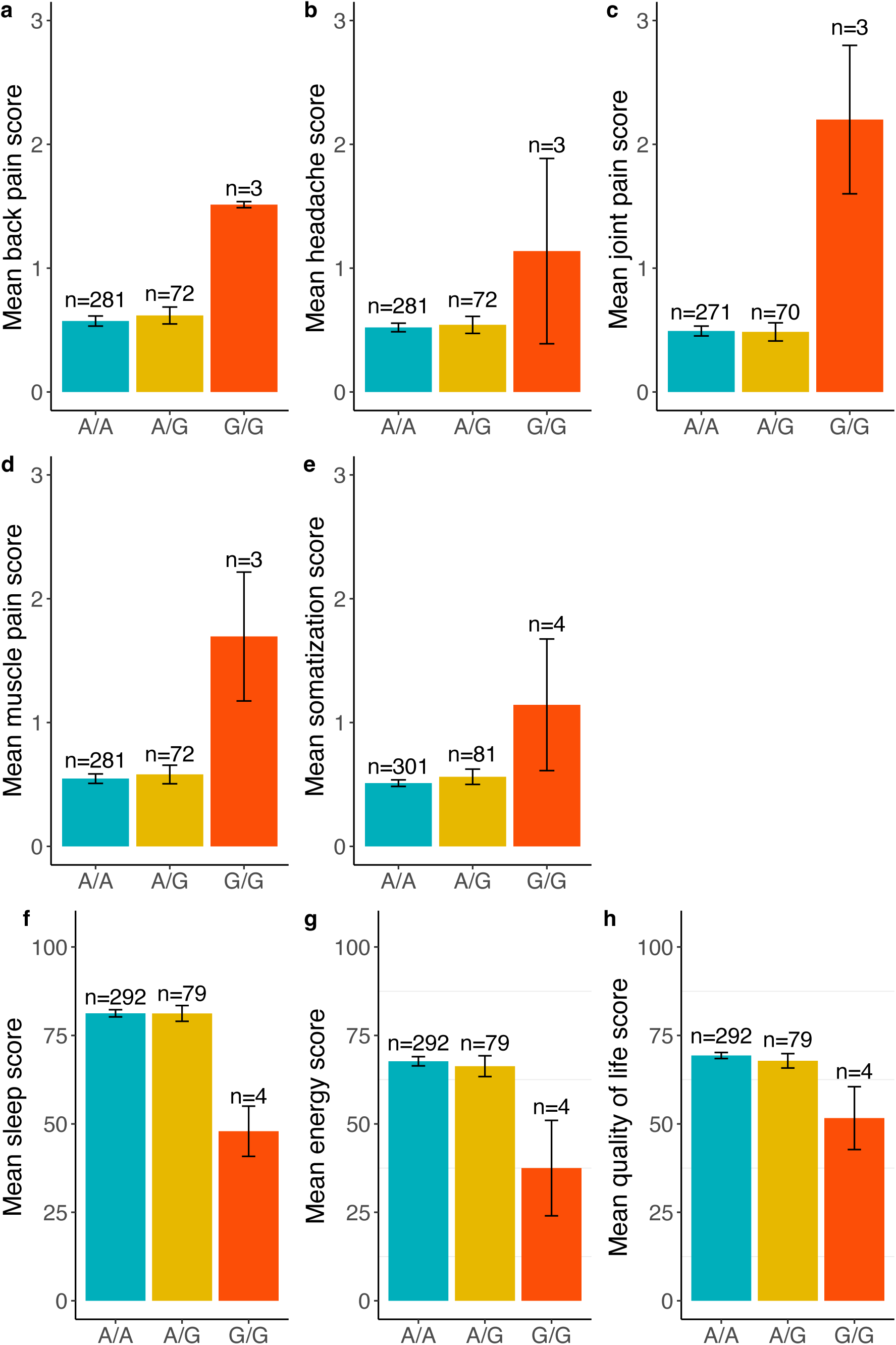
Association of rs2013566 with symptom traits in individuals with IBS. In these bar graphs, the x-axis represents the genotypes at rs2013566, while the y-axis represents the mean scores of each trait (as indicated below). Each bar represents a different genotype group. Error bars extending from each bar indicate the standard error (SE) from the mean. The traits represented by the labels are as follows: (a) back pain; (b) headache; (c) joint pain; (d) muscle pain; (e) somatization; (f) sleep; (g) energy levels; (h) quality of life.

**Figure 4.**
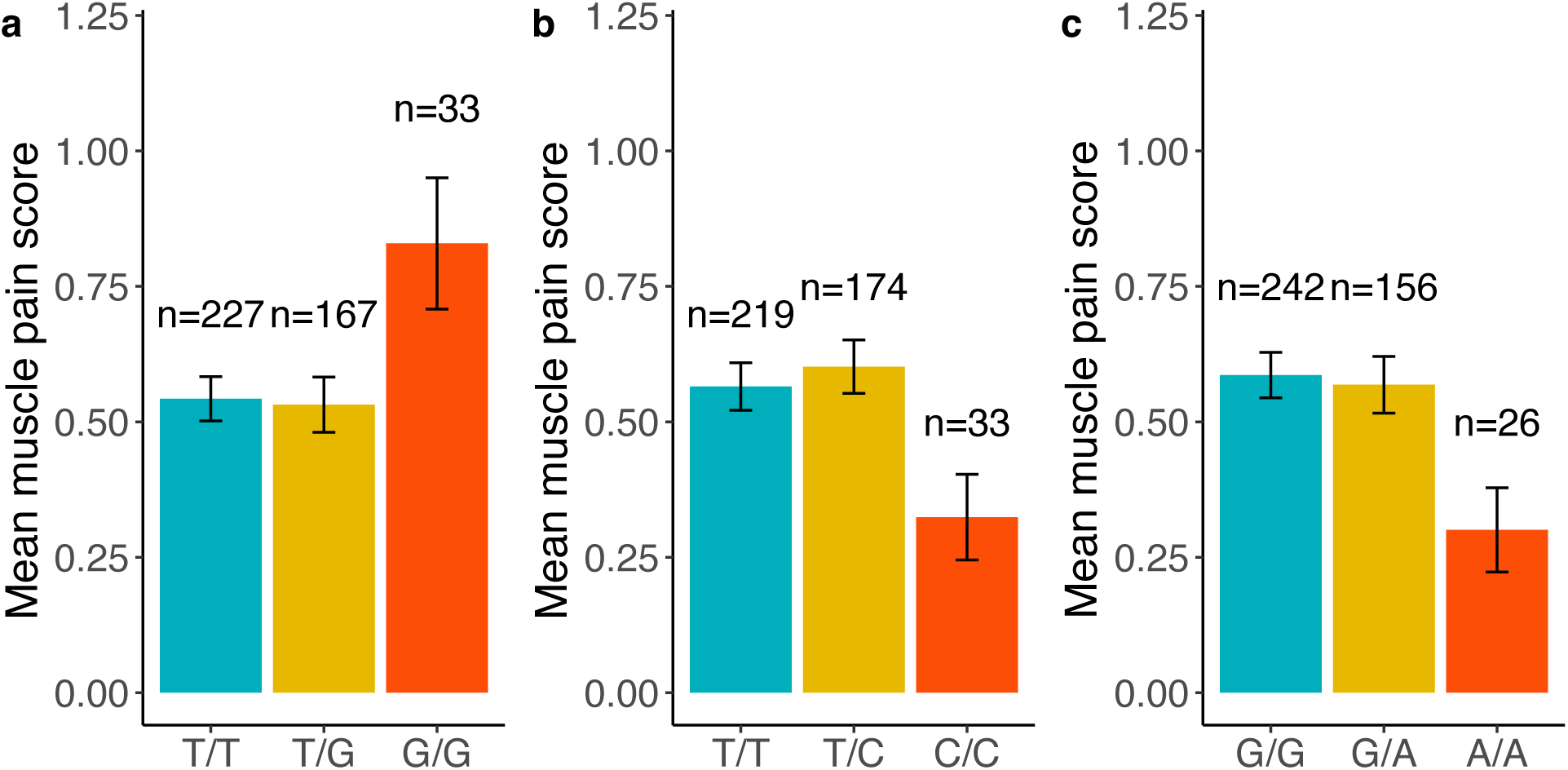
Associations of other SNPs in *NTRK2* with muscle pain in participants with IBS. These bar graphs display the associations between different SNPs in NTRK2 and muscle pain in participants with IBS. The specific SNPs in this figure include: (a) rs1147198; (b) rs1627784; (c) rs1624327.

**Table 4.** Significant associations between SNPs and IBS-related symptoms in primary and UKBB data.

| Symptoms | Primary Data |  |  |  |  | ImpUKB |  |  |  | ES200K |  |  |  |
| --- | --- | --- | --- | --- | --- | --- | --- | --- | --- | --- | --- | --- | --- |
| | SNP ID | R/A | n | $\beta$ (SE) | *p* | n | OR | 95% CI | *p* | n | OR | 95% CI | *p* |
| **Symptom domain i. Abdominal pain and other somatic symptoms** |  |  |  |  |  |  |  |  |  |  |  |  |  |
| Headache | rs2013566 | A/G | 356 | 0.37 (0.17) | 0.029 | 7476 | 1.11 | 0.93-1.33 | 0.24 | 3060 | **1.34** | 1.03-1.74 | **0.028** |
| Back pain | rs2013566 | A/G | 356 | 0.49 (0.19) | 0.011 | 7474 | **0.79** | 0.65-0.95 | **0.01** | 3055 | 1.02 | 0.78-1.34 | 0.894 |
| Joint pain | rs2013566 | A/G | 344 | 0.83 (0.17) | <0.001 | 7445 | 1.02 | 0.82-1.28 | 0.85 | 2995 | 1.19 | 0.85-1.67 | 0.313 |
| Somatization | rs2013566 | A/G | 386 | 0.32 (0.13) | 0.012 | 861 | 0.83 | 0.35-1.98 | 0.67 | 336 | 0.69 | 0.13-3.57 | 0.654 |
| Muscle pain | rs2013566 | A/G | 356 | 0.57 (0.18) | 0.002 | 1176 | 0.92 | 0.47-1.81 | 0.81 | 492 | 1.34 | 0.57-3.15 | 0.507 |
|  | rs1147198* | T/G | 427 | 0.14 (0.06) | 0.015 | 1176 | 1.41 | 0.93-2.12 | 0.1 |  |  |  |  |
|  | rs1627784 | T/C | 426 | -0.12 (0.06) | 0.032 | 1176 | 1.09 | 0.73-1.61 | 0.68 | 492 | 1.29 | 0.73-2.27 | 0.382 |
|  | rs1624327 | G/A | 424 | -0.14 (0.06) | 0.034 | 1176 | 1.35 | 0.91-2.01 | 0.14 | 492 | 1.42 | 0.81-2.52 | 0.223 |
| **Symptom domain iii. IBS-related quality of life** |  |  |  |  |  |  |  |  |  |  |  |  |  |
| Overall QoL** | rs2013566 | A/G | 375 | -8.85 (3.93) | 0.025 |  |  |  |  |  |  |  |  |
| Sleep | rs2013566 | A/G | 375 | -16.41 (4.49) | <0.001 | 1134 | 0.91 |  | 0.78 | 399 | 1.17 |  | 0.782 |
| Energy | rs2013566 | A/G | 375 | -15.49 (5.88) | 0.009 | 4458 | 0.88 |  | 0.49 | 1581 | 1.15 |  | 0.598 |

\* SNP not genotyped for muscle pain in ES 200K cohort
\*\* Trait does not present in the UKB Imp cohort
No data available in greyed cells.
CI: confidence interval, ES200k: UKBB participants with whole exome sequencing for comprehensive genetic variant identification, ImpUKB: imputed genotypes of UKBB participants using Haplotype Reference Consortium (HRC) and UK10K haplotype resources, n: number of sample, OR: odds ratio, *p*: *p*-value, QoL: quality of life, R/A: reference/alternative alleles, SE: standard error, SNP: single nucleotide polymorphism, UKBB: United Kingdom Biobank, $\beta$ : beta-coefficient relative to the alternative allele.

### External validation using UKBB data revealed that rs2013566 is associated with other traits

To validate our primary findings, we conducted an external validation using independent panels of UKBB data, including ES200k and ImpUKB. Among our 11 candidate SNPs, six (rs2013566, rs1627784, rs1624327, rs1047896, rs1187286, and rs41277883) showed significant associations with various somatic and psychological symptoms in individuals with IBS in the UKBB data (p < 0.05, Supplemental Table 4). Among these six SNPs, three (rs2013566, rs1627784, and rs1624327) were also identified as significant in our primary analysis. Importantly, the findings of these three overlapping SNPs either supported or contradicted the validation results from UKBB data. Specifically, the minor allele of rs2013566 consistently increased the likelihood of headaches (β = 0.37, p = 0.03), not only in our U.S. study population but also among individuals with IBS in the U.K. in the ES200k dataset (odds ratio [OR] = 1.34, 95% CI = 1.03-1.74; Table 4). However, it exhibited contrasting associations with back pain, being positively associated in our primary data (β = 0.49, p = 0.01), but negatively associated in the ImpUKB (OR = 0.79, 95% CI = 0.65-0.95; Table 4). In our primary analysis, we observed a negative association between the minor allele of rs1627784 and muscle pain (β = -0.12, p = 0.03). Although this association was not significant in both UKBB datasets, a distinct association with increased odds of depression (OR = 1.58, 95% CI = 1.11-2.24) was identified. Similarly, the association between the minor allele of rs1624327 and reduced muscle pain (β = -0.14, p = 0.03) was not supported by both UKBB datasets. Instead, rs1624327 was associated with decreased somatization (OR = 0.53, 95% CI = 0.3-0.91) and increased depression (OR = 1.57, 95% CI = 1.1-2.24) in the ImpUKB dataset, as well as increased headache (OR = 1.27, 95% CI = 1.07-1.5) in the ES200k dataset. Overall, the external validation not only confirmed the association between rs2013566 and an increased risk of headaches but also revealed a complex relationship between these SNPs and various somatic and psychological symptoms in individuals with IBS, which may vary across specific populations.

## Discussion

Although there is growing recognition of the impact of genetic predisposition on physio-psychological symptoms and quality of life in IBS, the role of genetic associations and their mechanistic contribution underlying these associations remain largely unexplored. This study aims to fill this gap by investigating the role of polymorphisms in the *BDNF/NTRK2* genes in contributing to comorbid symptoms in people with IBS. By examining the association between SNPs in the *NTRK2* and *BDNF* genes with a range of symptom traits, we discovered that several SNPs are significantly associated with the manifestation of various somatic symptoms and quality of life. Notably, individuals with a homozygous recessive genotype (G/G) at rs2013566 showed positive associations with multiple somatic symptoms and negative associations with quality of life. The association between rs2013566 and headaches in individuals with IBS was further validated by the UKBB data. Additionally, we identified three other SNPs that were associated with muscle pain and reduced quality of life. These findings provide insights into the complex genetic factors that may underlie the diverse symptoms experienced by individuals with IBS, which could provide valuable insights for the design of future research studies and for clinical intervention.

The BDNF/TrkB signaling pathway plays a critical role in the development, survival, plasticity, and functioning of neurons in the central nervous system (CNS), and its effects vary depending on cell type, tissue, age, and context [29]. The TrkB gene (*NTRK2*) is highly expressed in the mammalian brain and other tissues, with notable alternative splicing primarily observed in the brain [51–54]. During CNS development, the predominant isoform is TrkB.FL, while TrkB.T1 expression gradually increases after birth and reaches peak expression in adulthood [55]. Interestingly, TrkB.T1 is the primary isoform outside the nervous system, found in the heart, kidney, lung, and pancreas [56–58]. Previous studies have shown that astrocytes in the CNS specifically express TrkB.T1 and exhibit significant upregulation of the receptor mRNA and protein coincident with nociception and neuropathic pain following spinal cord injury in rodents [20, 21, 59]. Given the predominant expression of TrkB.T1 in the adult brain and its upregulation associated with pain, our observed association between rs2013566 and headaches in individuals with IBS is biologically plausible. The contribution of TrkB.T1 signaling to pain involves various potential mechanisms, including the mediation of inositol-1,4,5-trisphosphate (IP3)– dependent calcium release from intracellular stores [56, 60, 61]. Notably, independent studies in mouse models have demonstrated that such calcium-mediated signaling and imbalances between TrkB.FL and TrkB.T1 isoforms impact motor function in amyotrophic lateral sclerosis [62] and cardiac contractility in cardiomyopathy [56]. These findings provide additional evidence supporting the genotypic association between rs2013566 and physiological functions and potential correlations with back pain, joint pain, muscle pain, and somatization identified in our study. This association may potentially be explained by TrkB.T1-dependent alterations in calcium homeostasis and signaling pathways involving IP3 receptors in non-brain tissues, which contribute to the sensitization of pain pathways, amplification of nociceptive signals, and the development of musculoskeletal pain in individuals with IBS.

The association between TrkB.T1 SNPs and a spectrum of pain manifestations can potentially be further explained by their capacity to function as enhancer elements. The genomic location of the majority of SNPs significantly associated with somatic and psychological symptoms within the 3’UTR indicates their potential as enhancer elements involved in the regulation of gene expression across diverse tissues. TrkB.T1, which shares common exons with TrkB.FL up to exon 15, has a distinct exon 16. This exon contains 33 nucleotides that encode a unique intracellular 11 amino acid domain, a stop codon, and a unique 3’UTR sequence with multiple polyadenylation sites [29]. Importantly, the 3’UTR of the TrkB.T1 isoform is distinct from that of the 3’UTR for TrkB.FL and other alternatively spliced isoforms of the gene. Combined epigenetic mechanisms, involving histone modifications (H3K4me1 and H3K27ac), DNA methylation, and microRNA binding, impact the regulation of TrkB.T1 expression. [63–65]. Histone modification markers and transcription factors, in particular, play a critical role in establishing cell-type-specific gene expression patterns within active chromatin regions [66, 67]. According to HaploReg v4.2 [68], rs2013566, rs1627784, and rs1624327 exhibit enhanced H3K4me1 and H3K27ac markers not only in various brain tissues but also in rectal mucosa-derived cells and fetal leg/truck muscle (Supplemental Table 5). Despite the limited availability of epigenetic data in non-brain tissues, the presence of H3K4me1 and H3K27ac markers near these SNPs suggests that they may be located in close proximity to enhancer elements, potentially influencing the regulation of TrkB.T1 gene expression in gastrointestinal and muscle tissues. Among these, rs2013566 ranks 4 in RegulomeDB, ranging from 1 (most likely functional) to 6 (least likely functional) [69], further supporting its potential enhancer function and suggesting a probable influence on transcription factor binding (Supplemental Table 5).

Since none of these SNPs are considered expression quantitative trait loci (eQTLs) in any tissues available in a public database (GTExPortal, https://www.gtexportal.org), their potential of functional activity in the TrkB.T1 signaling is anticipated to be indirect rather than direct. Therefore, the identification of rs2013566 as potentially exhibiting enhancer regulatory activity in the brain, muscular, and gastrointestinal tissues supports the functional relevance of the observed associations between this SNP and a range of symptoms, particularly headache and musculoskeletal pain, in individuals with IBS. The remaining SNPs, rs1627784, and rs1624327, demonstrate relatively limited evidence of regulatory function, emphasizing the importance of further investigation to elucidate their potential significance.

Besides our primary analysis, we discovered a new and significant association between the number of minor alleles at rs41277883 and a 3.4-fold increased likelihood of having IBS. Interestingly, despite this association, this SNP is not directly associated with any of the somatic or psychological symptoms commonly observed in individuals with IBS. Nevertheless, rs41277883, located in the 3’UTR, overlaps with evolutionarily conserved elements found in the aforementioned SNPs, suggesting its potential role as an enhancer (Supplemental Table 5). A previous study associated rs41277883 with total pain intensity in individuals aged 65 years or older who underwent surgical repair for a hip fracture [70]. Considering these findings, the association between rs41277883 and the risk of IBS indicates its potential involvement in mechanisms related to pain sensitivity. Although these findings suggest that rs41277883 may serve as a potential genetic marker associated with an increased likelihood of developing IBS, further research is necessary to fully comprehend the implications of this association and to uncover the specific mechanisms by which rs41277883 influences the development and manifestation of IBS.

Conversely, the *BNDF* SNP rs6265 is widely recognized for its role in the substitution of valine (Val) with methionine (Met), leading to impaired functioning of processes involved in regulating extracellular BDNF levels [71]. Previous studies have extensively explored the genetic impact of rs6265, encompassing not only pain but also psychological and other somatic pain traits that are highly prevalent in individuals with IBS, including migraines [72], fibromyalgic pain [73], menstrual pain [74], anxiety [75, 76], and depression [77, 78]. In the context of IBS, our previous research revealed decreased plasma BDNF levels in IBS patients compared to non-IBS individuals [79]. Other studies have also found an association between lower serum BDNF levels and higher levels of anxiety and depressive symptoms in individuals with IBS [80]. In contrast, preclinical evidence suggests that higher amounts of BDNF and TrkB proteins were detected in the thoracolumbar spinal cord of IBS-like rats compared to control rats, and the administration of a selective TrkB antagonist reduced their visceral hypersensitivity [81]. Interestingly, despite its association with various conditions, rs6265 is not directly linked to any of the symptom traits observed in our analysis. This suggests that TrkB signaling, and the expression levels of the TrkB.T1 receptor, rather than extracellular BDNF concentration, may play a more significant role in determining the somatic or psychological symptoms reported in IBS. Alternatively, there is a possibility that the BDNF/TrkB response varies across tissues, or that the manifestation of symptoms is the result of multiple interacting factors.

Our findings also suggest that the genetic variations identified in the *NTRK2* gene may have implications for the occurrence of a wide range of somatic and psychological symptoms experienced by individuals with IBS, potentially impacting their quality of life. The variability in scores across the three symptom domains indicates differing levels of impairment among individuals with IBS, necessitating personalized approaches to symptom management and improving their quality of life. A deeper understanding of genetic predisposition can aid in tailoring symptom management strategies. In our investigation, we identify that individuals with IBS who carry a homozygous recessive genotype (G/G) at rs2013566 are more likely to experience headaches and other somatic pains, sleep disturbances, and reduced energy levels. Such genetic association underscores the importance of understanding how the homozygous recessive genotype at rs2013566 is linked to various somatic pain manifestations, as it can contribute to sleep disturbances and hinder energy restoration, ultimately leading to a decline in overall quality of life. Having the genetic information at the outset of the diagnosis could help healthcare providers to be aware of the risk for the above IBS phenotypes and intervene early to potentially prevent them. However, further investigation is required to comprehensively interpret these associations and patterns.

This study has several limitations that should be acknowledged. First, our focus was limited to specific SNPs within the *NTRK2* and *BDNF* loci, as we hypothesized that SNPs within the regulatory region of the *NTRK2* gene and/or the *BNDF* Val66Met SNP could impact on our phenotype of interest. Thus, there may be additional SNPs within these genes or related genes that could also contribute to the phenotype. The absence of an examination of susceptibility alleles at multiple loci also raises the possibility that unexplored SNPs may also contribute to the development of symptoms among those with IBS. Additionally, the failure to genotype certain SNPs (rs2013566 and rs3654) in one of the five studies may have introduced biases by missing crucial data for the interpretation of genetic associations with any of these symptoms. To address these limitations, future investigations should prioritize comprehensive genotyping in all relevant samples to ensure accurate results and enable a comprehensive interpretation of genetic associations with symptoms in those with IBS. Second, the study addressed potential bias arising from disparities in marital status and family income between the IBS and HC groups. These demographic factors could confound the study results by impacting the experience and management of IBS-related health conditions, as well as epigenetic signatures. To mitigate the impact of potential bias, interventions, and Hawthorne effects, only baseline data from all five studies were analyzed, and all regression models were adjusted for age, sex, and study design. Third, although environmental and lifestyle factors were not the primary focus of this study, they can influence the development and manifestation of these symptoms and should be considered in future studies. Finally, the recruitment of participants from a specific geographical area in the U.S. and the validation of primary findings using data from the U.K. limit the generalizability of the study to the broader population of individuals with IBS, urging caution in applying the results to diverse ethnicities or geographical regions.

In summary, our study identified a genetic association between TrkB.T1 SNPs, particularly rs2013566, and symptoms such as headache, musculoskeletal pain, and impaired quality of life in individuals with IBS. Moreover, we observed regulatory potential of these SNPs in relation to somatic and psychological symptoms. These findings suggest the potential for personalized treatment approaches and targeted symptom management based on genetic markers, in conjunction with the further understanding of somatic-psychological and environmental influences. Further research, including large-scale longitudinal observational studies, clinical trials, and exploration of underlying neurogenetic mechanisms, is necessary to gain a comprehensive understanding of these genotypic associations.

## Data Availability

In adherence to ethical and privacy standards, we are unable to deposit individual genotype data, while we remain committed to promoting research transparency within regulatory guidelines.

## Acknowledgements

The primary data sources were supported by the National Institute of Nursing Research grants: R01NR004101; R01NR004142; R01NR014479; R01NR01094; P30NR004001, as well as K23NR020044 for KK and T32NR016913 for HH. The content is solely the responsibility of the authors and does not necessarily represent the official views of the National Institutes of Health. The validation portion of this research was conducted using the UK Biobank Resource under Application Number 49852. The authors wish to thank Ernest Tolentino for the processing of the samples.

